# Road environment characteristics and fatal crash injury during the rush hour period in the U.S.: Model testing and nested analytical study

**DOI:** 10.1101/2022.06.09.22276199

**Authors:** Oluwaseun Adeyemi, Rajib Paul, Eric Delmelle, Charles DiMaggio, Ahmed Arif

**Author notes:** **Corresponding Author** Oluwaseun J. Adeyemi, Department of Public Health Sciences, University of North Carolina at Charlotte.

## Abstract

**Background:** A substantial proportion of crash injuries occur during the rush-hour period. This study aims to assess the relationship between county-level road environmental characteristics and fatal road crash counts during the rush-hour period.

**Method:** We merged eight-year (2010 - 2017) data from the Fatality Analysis Reporting System. We limited the data to crashes during the rush hour period (6–9 am; 3– 7 pm). The outcome variable was the counts of fatal crashes per county. The predictor variables were road design (intersection, driveway, ramp, work-zone), road type (interstate, highways, roads/streets), and inclement weather factors (rain, fog, snow). A nested spatial negative binomial regression model was used to estimate the rate ratio of fatal crash injury during the rush-hour period, with estimated county population sizes used as the offset variable. Small area estimates, adjusted crash fatality rates, clusters, and outliers were visualized using choropleths maps.

**Results:** The median prevalence of rush-hour-related fatal crashes was 7.3 per 100,000 population. Case-specific fatality rates from interstates, highways, roads, streets, intersections, rain, fog, and snow were higher than the median fatality rates. Also, the median crash fatality rates were significantly higher in rural counties as compared to urban counties. During the rush-hour period, fatal crash injury rates were significantly elevated on interstates, highways, roads and streets, intersections, driveways, and work zones. Further, rain and fog were significantly associated with elevated fatal crash rates during the rush-hour period. Spatial clusters of fatal crash injuries were found in counties located in Idaho, Montana, Nevada, California, Wyoming, Utah, and across a few states in the Southeast.

**Conclusion:** Certain built, and natural road environment factors may influence crash injury rates during the rush-hour period.

## 1. Introduction

Road crashes are preventable causes of morbidity and mortality in the United States (U.S.). In 2017, there were 6.5 million crashes, which accounts for the death of over 37,000 individuals and about 2.8 million injuries. One person dies every 14 minutes from crash-related events each day in the U.S. (National Center for Statistics and Analysis, 2019a). In 2016, over 2.5 million individuals were treated for crash-related injuries in the emergency departments across the US (National Center for Statistics and Analysis, 2017a). The cost of health care and loss from productivity exceeded 75 billion dollars in 2015 (Center for Disease Control and Prevention, 2020a). Fatal and non-fatal crashes are disproportionately distributed across the day, with crash injuries predominantly occurring around the rush-hour period (Varghese & Shankar, 2007).

The rush-hour period represents the time of the day in which the roads have the highest densities of human and automobile activities (Call, Medina, & Black, 2019; Norros, Kuusela, Innamaa, Pilli-Sihvola, & Rajamaki, 2016). In the U.S., the rush hour is between 6 to 10 am and 3 and 8 pm (Call, Wilson, & Shourd, 2018; Paleti, Eluru, & Bhat, 2010; Xu & Xu, 2020). This period varies by county and rurality (Jaffe, 2014), with urban communities in North Carolina, for example, having one of every four road crashes occurring during the rush hour (Tippett, 2014). The evening rush hour witnesses more crash events than the morning rush hour period (HG.org, 2020; Tippett, 2014; Varghese & Shankar, 2007).

## 2. Background

An individual’s geographical location is an important social determinant of health (González, Wilson-Frederick Wilson, & Thorpe, 2015; Healthy People, 2020), and the road environment has long been associated with fatal and non-fatal crashes injuries (National Highway Traffic Safety Administration, 2010). In the U.S., over 1.2 million fatal and non-fatal crash injuries occur at or near intersections (Federal Highway Administration, 2020b; National Highway Traffic Safety Administration, 2010). Factors reported to be associated with intersection-related crash injuries include driving inattention, misjudgment of the speed of another vehicle, distracted driving, and aggressive driving (Federal Highway Administration, 2020b; National Highway Traffic Safety Administration, 2010). Inadequate surveillance is associated with a six-fold increased crash risk at intersections compared to non-intersections, while misjudgment of another vehicle’s speed is associated with a four-fold increased risk of intersection-related crash events (National Highway Traffic Safety Administration, 2010).

While intersections represent an area where two or more roadways meet, driveways represent road stretches that lead into public or commercial roads (Liu, 2007; Nadler, Courcoulas, Gardner, & Ford, 2001). Driveways crash injuries commonly involve slow-moving vehicles, backup driving, crashes from making a left or right turn into a major road (Liu, 2007; Nadler et al., 2001). Child pedestrians are commonly involved in drive-way-related crash injuries (Anthikkat, Page, & Barker, 2013; Nadler et al., 2001). A systematic review reported that residential driveways are associated with over three-fold crash injury (Anthikkat et al., 2013). Also, driveways that exit into a local road have longer lengths and run along property boundaries are associated with a three-to-five-fold increased risk of crash injury (Anthikkat et al., 2013; Shepherd, Austin, & Chambers, 2010).

Work zones represent non-permanent road characteristics that have been associated with property damage and crash injury (American Road & Transportation Builders Association, 2018; Federal Highway Administration, 2019). In the U.S., about five percent of all fatal crashes occur at work zones (American Road & Transportation Builders Association, 2018). Work zone-related crash events increased from 84,000 in 2009 to 123,000 across the U.S., with the associated injuries rising from 19,000 to 31,000 (American Road & Transportation Builders Association, 2018). It is estimated that one work zone fatal injury occurs every four billion vehicle mile traveled (VMT) (Federal Highway Administration, 2019). Also, the work zone area length and the frequency of work zone regions are associated with increased crash injury and property damage (Chen & Tarko, 2012; Ozturk, Ozbay, & Yang, 2014; Athanasios Theofilatos, Ziakopoulos, Papadimitriou, Yannis, & Diamandouros, 2017).

Earlier studies have identified an increased risk of crash injuries during the rain (Andrey & Yagar, 1993; Jung, Jang, Yoon, & Kang, 2014; Qiu & Nixon, 2008; A. Theofilatos & Yannis, 2014), snow (El-Basyouny, Barua, & Islam, 2014; Fridstrøm, Ifver, Ingebrigtsen, Kulmala, & Thomsen, 1995; A. Theofilatos & Yannis, 2014), and fog (A. Theofilatos & Yannis, 2014; Wu, Abdel-Aty, & Lee, 2018). Between 2007 and 2016, about 8% of fatal crash injuries were associated with rain, while fog and snow were each related to 2% of all fatal crash injuries across the U.S. (Federal Highway Administration, 2020a). Rain, fog, and snow each accounted for 46%, 9%, and 10%, respectively, of weather-related fatal crash counts (Federal Highway Administration, 2020a). About 300 - 400 fog-related fatal crashes occur yearly in the U.S. (Hamilton, Tefft, Arnold, & Grabowski, 2014; Wu et al., 2018), and snow accounts for 16% of all weather-related crashes (Federal Highway Administration, 2020a). The conceptual link between these inclement weather factors and fatal crash events is related to driving visibility and road surface friction. Rain, fog, and snow are associated with low visibility, while rain and snow limit skid resistance due to reduced surface friction (El-Basyouny et al., 2014; Li et al., 2019).

Central to the built and natural environmental characteristics of the crash scene is the rural-urban status. The U.S. rural community is home to about 20 percent of the U.S. population (United States Census Bureau, 2019a), and less than a third of vehicle miles traveled in the U.S. occurs in the rural areas (Federal Highway Administration, 2018; Insurance Institute for Highway Safety, 2019). However, rural counties have about half of all fatal crashes (Insurance Institute for Highway Safety, 2019; National Center for Statistics and Analysis, 2019b). Earlier studies have reported speeding as a major risky driving behavior that contributes to fatal crash injuries in rural communities (Federal Highway Administration, 2000b; Insurance Institute for Highway Safety, 2019). However, the rural areas have poorer road qualities, evidenced by an increased proportion of structurally deficient bridges and poor pavement conditions (Congressional Research Service, 2018). Additionally, rural communities have fewer hospitals and health-related infrastructures (Center for Disease Control and Prevention, 2020c; Pink, Osgood, & Sana, 2020) and longer response time (Byrne et al., 2019; King, Pigman, Huling, & Hanson, 2018; K. E. M. Miller, James, Holmes, & Van Houtven, 2020). With a larger proportion of the older population living in the rural community (Smith & Trevelyan, 2019; United States Department of Agriculture, 2019), the risk of fatal injury is further heightened with the increased prevalence of co-morbid conditions and disabilities (Garcia et al., 2019; Zhao, Okoro, Hsia, Garvin, & Town, 2019).

In predicting crash occurrence at the county level, there is a need to establish the independence of observations to reduce analytical errors (Sainani, 2010). The possibility exists that the occurrence of crash events in a county may increase (positive autocorrelation) or reduce (negative autocorrelation) the likelihood of its occurrence in neighboring counties, especially, if such counties share similar exposures – a concept defined as spatial autocorrelation (Kirby, Delmelle, & Eberth, 2017). Spatial autocorrelation techniques, commonly with the use of global Moran’s I (Anselin, 1988) or general Getis-Ord (Getis & Ord, 2010), presents ways for adjustings for spatial autocorrelation. Additionally, crash events share unobserved roadway characteristics (Carson & Mannering, 2001), and these spatial estimators adjust for the unobserved environmental elements (Jonathan, Wu, & Donnell, 2016). Earlier studies have suggested including spatial estimators in crash injury risk modeling (Jonathan et al., 2016; Lord, Cloutier, Garnier, & Christoforou, 2018). According to Tobler’s first law of geography (Sui, 2004), all things are related, but close things are more related than far things. With increasing distance, the global Moran’s I value tends to reduce (Epperson, 2005). Detecting spatial clusters, therefore, rely on using local estimators of spatial autocorrelation (Waller & Gotway, 2004) such as local Moran’s I (Anselin, 1995) and Getis-Ord GI* (Getis & Ord, 2010; Ord & Getis, 1995). Additionally, county-level estimates may be better predicted using small area estimation techniques, and the spatial structure of counties provides more accurate estimation compared to national or state-level estimates (Kirby et al., 2017).

Identifying the environmental factors associated with fatal road crashes and their spatial distribution is important to create focused intervention and resource allocation. It is unknown to what extent road types, road designs, and inclement weather conditions associates with fatal crash events within the rush hour period. To our knowledge, no publicly available study reported fatal crash injury rates during the rush hour period. The literature on rush hour-related crash injury is sparse, and this study seeks to provide substantial information on the environmental factors associated with fatal crash events during the rush hour period. Therefore, this study aims to assess the relationship between county-level road environmental characteristics and fatal road crash rates. It is hypothesized that county-level measures of road types (such as interstate, highways, roads, and local streets), road designs (such as intersections, driveways, ramps, and work zones), and the natural environment (such as rain, snow, and fog) will be associated with increased rates of fatal crash events. Additionally, this study aims to identify clusters of fatal crash injuries during the rush-hour period. It is hypothesized that homogenous clusters of fatal crash events will emerge from predicted estimates of the county-level fatal crash rates.

## 3. Methods

### 3.1 Study Design

This ecological study pooled eight years of data (2010 – 2017) from the Fatality Analysis Reporting System (FARS). The FARS dataset is a repository of fatal road crash events hosted by the National Highway Traffic Safety Agency (NHTSA). It provides a nationwide census of all crash injuries involving at least a fatality across counties in the U.S. and the District of Columbia (National Highway Traffic Safety Administration, 2017). Data are released every year in multiple linkable files across domains representing the crash scene, person-related, and vehicle-related information (National Highway Traffic Safety Administration, 2016a). For this study, the variables were extracted from the accident file.

### 3.2 Inclusion and Exclusion Criteria

This study’s inclusion criterion was that the crash event must have occurred during the rush hour period. We defined the rush hour crashes as road accidents that occurred between 6 to 10 am and 3 to 7 pm (Adeyemi, Arif, & Paul, 2021; Federal Highway Administration, 2017). We restricted the data to counties within the conterminous U.S., excluding counties in Alaska, Hawaii, Northern Mariana Islands, U.S. Virgin Islands, American Samoa, Guam, and Puerto Rico. Each county was classified as either urban or rural using the Rural-Urban Commuting Area (RUCA) code (Economic Research Services, 2019). Counties that were classified within the range of metropolitan to high commuting micropolitan were classified as urban, while low commuting micropolitan to rural areas were classified as rural. The final data consisted of 3,102 counties, with 1,691 classified as urban while 1,411 classified as rural (Figure 1).

**Figure 1:**
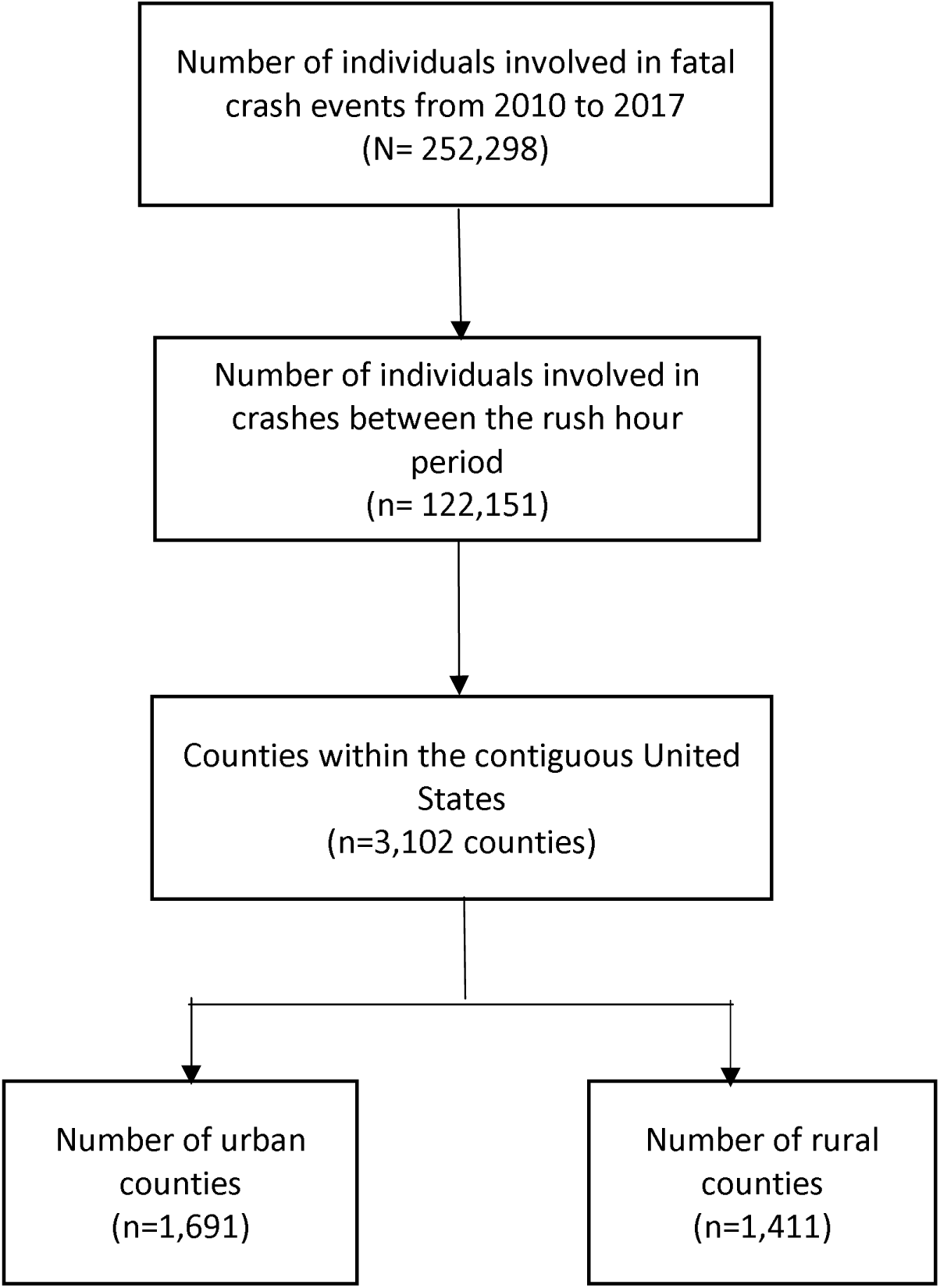
Data selection and aggregation steps

### 3.3 Data Processing

The data extracted from the accident file included the state and county codes, the year and hour of the crash, the route the collision occurred (route-related accidents), the relationship of the crash to a junction or a specific location (junction-related accidents), the relationship of the accident to the boundaries of work zones (work zone accident), atmospheric conditions (weather-related accidents), and the number of fatalities that occurred with each crash. The raw data file reported route-related crashes as a single variable with multiple categories comprising interstate roads, U.S. highway, state highway, county road, local streets in townships, municipality, and frontage other roads, and unknown. Three dummy road type variables were generated from this variable: interstate, highways (U.S. highway + state highway), and roads and streets (county road + local streets in townships, municipality, and frontage roads).

Similarly, junction-related crashes were coded originally as a multi-categorical nominal variable comprising of non-junction, intersection and intersection-related, driveway and driveway-related, ramp and ramp-related, railway grade crossing, crossover-related, shared-use path crossing, acceleration/deceleration lane, through the roadway, other location, unknown and not reported. Three road design dummy variables were generated from this variable: intersection (intersection + intersection-related), driveway (driveway + driveway-related), and ramp (ramp + ramp-related). Additionally, weather-related crashes were coded as multi-categorical nominal variables. These variables were weather (first weather condition that affects visibility), weather1 (second weather condition that affects visibility), and weather2 (any other weather condition that affects visibility). Each of these variables was reported in multiple categories: clear weather, no additional atmospheric condition, rain, sleet, snow, fog/smog/smoke, severe crosswinds, blowing sand/soil/dirt, blowing snow, freezing rain/drizzle, others, unknown, or not reported. Three inclement weather-related crashes dummy variables were generated across the weather variables: rain (rain + freezing rain/drizzle), snow (snow + blowing snow + sleet), and fog (fog/smog/smoke). Work zone-related crashes were reported as a multi-categorical nominal variable: None, construction, maintenance, utility, work zone type unknown, and not reported. This variable was re-categorized into a dummy variable: work zone (construction, maintenance, utility, type unknown). The number of fatalities per crash was measured as a continuous variable.

Data files across 2010 and 2017 were appended. A five-digit county Federal Information Processing Standard (FIPS) code was generated by concatenating the two-digit state code and the three-digit county codes.

### 3.4 Variable Definition

The outcome variable was the median fatal counts per county. The choice of using the median was based on the finding that the yearly distribution of the fatal counts per county was not normally distributed. To obtain the median fatal counts, the fatal counts for each county were aggregated by year to generate the yearly counts. Then, the median count across the years was computed. The average of the 2010 to 2017 county population estimates was used as the offset variable. The dummy variables were aggregated per county across the years. Counts of 0 represented an absence of the category of interest, while values of 1 and higher represented the presence of the category of interest in the county. The recoded dummy variables served as the predictor variables.

For this study, the county characteristics of interest that served as the control variables were the percentage of the White and male population, county rates of hospital utilization, unemployment, vehicle density, county gross domestic product (GDP), and median household income. Data on the county population, the percentage of the white and male population, median household income, and vehicle estimate per county were computed as the average of 2010 to 2017 estimate from the American Community Survey (United States Census Bureau, 2019b). The hospital utilization rate per county was obtained from the mean emergency department utilization per 1000 by Medicare beneficiaries per county (Center for Medicare and Medicaid Services, 2019). The mean unemployment rate per county was obtained from the 2010-2017 local area unemployment statistics of the U.S. Bureau of Labor (U.S. Bureau of Labor Statistics, 2019).

We created a spatial weight matrix of all the eligible county and county-equivalents in the dataset using the queen contiguity setting. The spatial weight matrix essentially maps the spatial relationship between the location, to establish the occurrence of spatial autocorrelation (Zhou & Lin, 2008). Spatial autocorrelation of the residuals provides information on the independence of the residuals. The presence of significant spatial autocorrelation suggests that there is a lack of independence of the residuals. We used the Euclidean distance with the k nearest neighbor set at 4.

### 3.5 Analysis

#### 3.2.1 Descriptive Statistics

We visualized the distribution of fatal counts per county. Further, we computed the rush hour-related raw and predicted median fatality rate by dividing the fatal counts per county by the county’s population estimates. Also, we reported the fatality rate specific to the road types (interstate, highway, road, and streets), road types (intersections, driveway, ramps, work zone), and inclement weather (rain, snow, fog). Differences in the crash-specific rates against their dummy variables were measured using the Mann-Whitney U test. We reported the distribution of rush hour fatality rate and the county characteristics across the urban and rural counties. Differences in the fatality rates and county characteristics were measured using independent T-tests and the Mann-Whitney U tests as appropriate.

#### 3.5.2 Regression Models

We used univariate negative binomial regression analysis to assess the relationship of all the predictor variables and county characteristics with fatal crash events. Variance inflation factor was used to assess multicollinearity.

We reported the adjusted incidence rate ratio for each of the ten variables in the road type, road design, and inclement weather group, adjusting for county characteristics. We then estimated a nested regression model using all determinants. Specifically, with road designs (intersection, driveway, ramp, and work zone) nested in road types (interstate, highways, and road/streets), interaction variables were generated for road design and road types, and these interaction variables were added to the model. After establishing evidence of spatial autocorrelation on all the regression models using the global Moran’s I, a corresponding nested spatial regression model was designed, and incidence rate ratios and the 95% confidence intervals (CI) were estimated. Spatial and non-spatial models were compared using the Akaike Information criteria (AIC) (Lee & Bell, 2009). Using Matérn covariance as a kernel function in the Gaussian process (Kammann & Wand, 2003), the final nested spatial regression models for each of the individual and all-determinant models are stated below:

##### Model 1: Interstate with nested road designs

#### *Y = β0_1_ + β1/nterstate + β2/nterstate * /ntersection + β3/nterstate * Driveway + β4Interstate * Ramp + β5Interstate * WorkZone + βγ_1_Covariates* + Matérn(1 | *longitude + latitude) + offset(log(popestimate)*

#### *Y = β0_2_+ β6Highway + β7Highway * Intersection + β8Highway * Driveway + β9Highway * Ramp + β10Highway * WorkZone + βγ_2_Covariates* + Matérn(1 | *longitude + latitude) + offset(log(popestimate)*

#### *Y = β0_3_+ β11Road + β12Road * Intersection + β13Road * Driveway + β14Road * Ramp + β15Road * WorkZone + βγ_3_Covariates* + Matérn(1 | *longitude + latitude) + offset(log(popestimate)*

#### *Y = β0_4_+ β16Rain + β17Fog + β18Snow + βγ_4_Covariates* +Matérn(1 | *longitude + latitude) + offset(log(popestimate)*

#### *Y = β0_5_+ β1Interstate + β2/nterstate * Intersection + β3Interstate * Driveway + β4/nterstate * Ramp + β5Interstate * WorkZone + β6Highway + β7Highway * Intersection + β8Highway * Driveway + β9Highway * Ramp + β10Highway * WorkZone + β11Road + β12Road * Intersection + β13Road * Driveway + β14Road * Ramp + β15Road * WorkZone + β16Rain + β17Fog + β18Snow + βγ_5_Covariates* + Matérn(1 | *longitude + latitude) + offset(log(population estimate)*

The all-determinant spatial model was the most parsimonious, and this model was used to generate small area estimates and the adjusted fatality rates per county. Cluster and outlier analysis was performed using Anselin’s local Moran’s to identify fatal crash events spatial clusters (Anselin, 1995). Also, a hotspot analysis was performed using the Getis-ORD star (Getis & Ord, 2010) to assess the spatial distribution of significant crash rates across neighboring counties.

Data analysis was performed using Stata version 16 (StataCorp, 2020) and R version 3.6.2 /R Studio version 1.2.5033 (R Core Team, 2019; RStudio Team, 2019). Specifically, the R-packages used for this study were the Spatial Dependence package (SPDEP) (Bivand et al., 2019), Modern Applied Statistics with S (MASS) (Ripley et al., 2019), and the Mixed-Effect Models, Particularly Spatial Models (spaMM) (Rousset, Ferdy, Courtiol, & GSL authors, 2020). Spatial weights and choropleths were created with ArcGIS Pro version 10.8 (Environmental Systems Research Institute, 2020).

## 4. Results

### 4.1. Fatal Injury Rates

Across the eight years, fatal road crashes were reported in 2,550 of the 3,102 counties. The median rush hour-related fatality rate per county was 7.30 (IQR: 11.1) per 100,000 population (Table 1). Across road types, the median (IQR) rush hour-related fatal crash injuries were highest on the highways (9.4 (10.8) per 100,000 population), followed by roads and streets (8.4 (9.7) per 100,000 population) and interstate (7.4 (9.9) per 100,000 population). Intersection-specific and driveway-specific median (IQR) fatal crash injuries were 7.8 (9.1) and 7.1 (7.8) per 100,000 population, respectively, during the rush hour period. The median (IQR) fog and rain-related fatal crash injuries during the rush hour period were 9.3 (10.5) and 7.8 (8.9) per 100,000 population, respectively, during rush hour period.

**Table 1:** Case-specific fatality rates from road environmental characteristics during the rush hour period between 2010-2017

| Categorical Variable | Median Fatal Rate (IQR)<br>(/100,000 population) |
| --- | --- |
| Road Type |  |
| Interstate-specific death | 7.4 (9.9) |
| Highway-specific death | 9.4 (10.8) |
| Roads and Streets- specific deaths | 8.4 (9.7) |
| Road Design |  |
| Intersection- specific deaths | 7.8 (9.1) |
| Driveway-specific deaths | 7.1 (7.8) |
| Ramp-specific deaths | 4.6 (3.4) |
| Work Zone-specific deaths | 6.7 (8.2) |
| Inclement * |  |
| Rain-specific deaths | 7.8 (8.9) |
| Fog-specific deaths | 9.3 (10.5) |
| Snow-specific deaths | 7.5 (9.0) |

There were significant differences in the median fatal crash injury rates by rural and urban status (p<0.001) (Table 2). The median (IQR) rush hour-related fatal crash injury rate in rural and urban counties was 9.5 (18.7) 6.3 (7.7) per 100,000 population. Other county characteristics that demonstrated significant rural-urban differences were the county-level hospital utilization (higher urban rates), unemployment rates (higher urban rates), household income (higher urban rates), the proportion of Whites (higher rural rates), males (higher rural rates), gross domestic product (higher urban rates), and vehicle density (higher rural rates).

**Table 2:** County characteristics and rush hour-related fatal crash injuries stratified by rural-urban status

| Variable (N=3102) | All counties<br>N=3,102 | Urban Counties<br>n=1,691 | Rural Counties<br>n=1,411 | p-value |
| --- | --- | --- | --- | --- |
| Median fatality rate (/100,000) | 7.3 (11.1) | 6.3 (7.7) | 9.5 (18.7) | <0.001<sup>b</sup> |
| Mean Hospital utilization per county | 702.8 (149.8) | 719.1 (132.1) | 683.5 (166.3) | <0.001<sup>a</sup> |
| Median Unemployment rate | 4.3 (1.8) | 4.4 (1.5) | 4.2 (2.3) | 0.001<sup>b</sup> |
| Median Household Income (/1000) | 48.7 (14.3) | 51.5 (16.7) | 46.2 (11.8) | <0.001<sup>b</sup> |
| Median % white population | 92.4 (15.2) | 89.0 (17.9) | 95.4 (8.1) | <0.001<sup>b</sup> |
| Mean % male population | 50.0 (2.2) | 49.6 (1.9) | 50.4 (2.4) | <0.001<sup>b</sup> |
| Median Average GDP (/100,000) | 7.9 (21.7) | 8.6 (24.8) | 7.0 (16.8) | 0.005<sup>b</sup> |
| Mean Vehicle density | 361.5 (38.3) | 357.3 (31.7) | 371.1 (43.0) | <0.001<sup>a</sup> |

a: Independent sample t-test b: Mann-Whitney U test; GDP: Gross Domestic Product

### 4.2 Rate Ratios of Fatal Injuries

In the univariate models, intersections, driveways, and ramps were associated with reduced rates of fatal crash injury across all counties during the rush hour period (Table 3). However, in rural counties, intersections (RR: 1.44; 95% CI: 1.30 - 1.60) and driveways (RR: 1.35; 95% CI: 1.13 - 1.62) were associated with 44% and 35% increased rates of fatal crash injuries, respectively. Also, across all counties, highways and roads and streets were associated with a two-fold (RR: 2.07; 95% CI: 1.89 - 2.28) and 9% (RR: 1.09; 95% CI: 1.02 - 1.17) increased rate of fatal crash injuries during the rush hour period. There was significantly elevated fatal crash injury in urban (RR: 1.86; 95% CI: 1.64 - 2.11) and rural highways (RR: 2.99; 95% CI: 2.65 - 3.38), in urban (RR: 1.11; 95% CI: 1.02 - 1.22) and rural (RR: 1.56; 95% CI: 1.41 - 1.73) roads and streets, and in rural interstate roads (RR: 1.85; 95% CI: 1.61 - 2.13). Across the rural counties, rain (RR: 1.41; 95% CI: 1.24 - 1.62), fog (RR: 1.52; 95% CI: 1.15 - 1.99), and snow (RR: 1.31; 95% CI: 1.07 - 1.59) were associated with significantly elevated fatal crash injury during the rush hour period.

**Table 3:** Negative binomial regression (non-nested) models assessing the unadjusted relationship between rush hour-related fatal road accidents and road environmental and county-level characteristics stratified by rural-urban status

| Variables | Univariate Models |  |  | VIF* |
| --- | --- | --- | --- | --- |
|  | All counties | Urban Counties | Rural Counties |  |
| **Road Design** |  |  |  |  |
| Intersections | **0.94 (0.88 – 0.99)** | 0.96 (0.89 – 1.04) | **1.44 (1.30 – 1.60)** | 1.57 |
| Driveways | **0.86 (0.80 – 0.94)** | 0.93 (0.85 – 1.01) | **1.35 (1.13 – 1.62)** | 1.23 |
| Ramps | **0.56 (0.50 – 0.63)** | **0.67 (0.60 – 0.75)** | 1.21 (0.40 – 3.85) | 1.31 |
| Work Zones | 0.94 (0.84 – 1.06) | 0.97 (0.86 – 1.10) | **1.69 (1.28 – 2.24)** | 1.10 |
| **Road Type** |  |  |  |  |
| Interstate | 0.96 (0.89 – 1.03) | 0.95 (0.88 – 1.03) | **1.85 (1.61 – 2.13)** | 1.31 |
| Highway | **2.07 (1.89 – 2.28)** | **1.86 (1.64 – 2.11)** | **2.99 (2.65 – 3.38)** | 1.36 |
| Roads and Streets | **1.09 (1.02 – 1.17)** | **1.11 (1.02 – 1.22)** | **1.56 (1.41 – 1.73)** | 1.33 |
| **Inclement Weather** |  |  |  |  |
| Rain | 0.95 (0.88 – 1.02) | 0.97 (0.89 – 1.05) | **1.41 (1.24 – 1.62)** | 1.29 |
| Fog | 1.23 (1.07 – 1.43) | 1.24 (1.07 – 1.46) | **1.52 (1.15 – 1.99)** | 1.03 |
| Snow | 0.89 (0.80 – 0.99) | 0.86 (0.78 – 0.96) | **1.31 (1.07 – 1.59)** | 1.08 |
| Hospital utilization | **1.00 (1.00 – 1.00)** | **1.00 (1.00 – 1.00)** | 0.99 (0.99 – 1.00) | 1.46 |
| Unemployment rate | **1.08 (1.05 – 1.10)** | **1.10 (1.07 – 1.13)** | 1.02 (0.99 – 1.05) | 1.48 |
| Household Income | **0.99 (0.99 – 0.99)** | **0.99 (0.99 – 0.99)** | **0.99 (0.99 – 0.99)** | 1.61 |
| % White population | **1.00 (1.00 – 1.00)** | **1.00 (1.00 – 1.00)** | **0.99 (0.99 – 0.99)** | 1.53 |
| % male population | **1.07 (1.05 – 1.09)** | **1.07 (1.04 – 1.09)** | **1.02 (1.00 – 1.05)** | 1.27 |
| Average GDP | **1.00 (1.00 – 1.00)** | **1.00 (1.00 – 1.00)** | 0.99 (0.99 – 1.00) | 1.01 |
| Vehicle density | 0.99 (0.99 – 1.00) | 0.99 (0.99 – 1.00) | **0.99 (0.99 – 0.99)** | 1.77 |

VIF: Variance Inflation Factor: values $\leq 3$ is accepted as this suggests no multicollinearity; Metropolitan status (VIF=1.48) was added to the final model as the model final model evidenced by a reduced AIC

After adjusting for county characteristics, intersections, interstate, highway, roads, and street, rain and fog were associated with significantly elevated fatal crash injuries during the rush hour period (Table 4). The adjusted spatial model showed that while intersection was associated with a 21% increased rate of fatal crash injuries (RR: 1.21; 95% CI: 1.13-1.28), ramps were associated with a 14% decreased rate of fatal crash injuries (RR: 0.84; 95% CI: 0.78-0.95). Also, the interstate (RR: 1.45; 95% CI: 1.32-1.59), highway (RR: 2.48; 95% CI: 2.25-2.72), and road/street (RR: 1.48; 95% CI: 1.37-1.60) were associated with increased rates of fatal crash injuries in the rush hour period. Rain (RR: 1.15; 95% CI: 1.08-1.23), fog (RR: 1.29; 95% CI: 1.15-1.47) and snow (RR: 1.15; 95% CI: 1.06-1.25) were each associated with increased fatal rates.

**Table 4:**
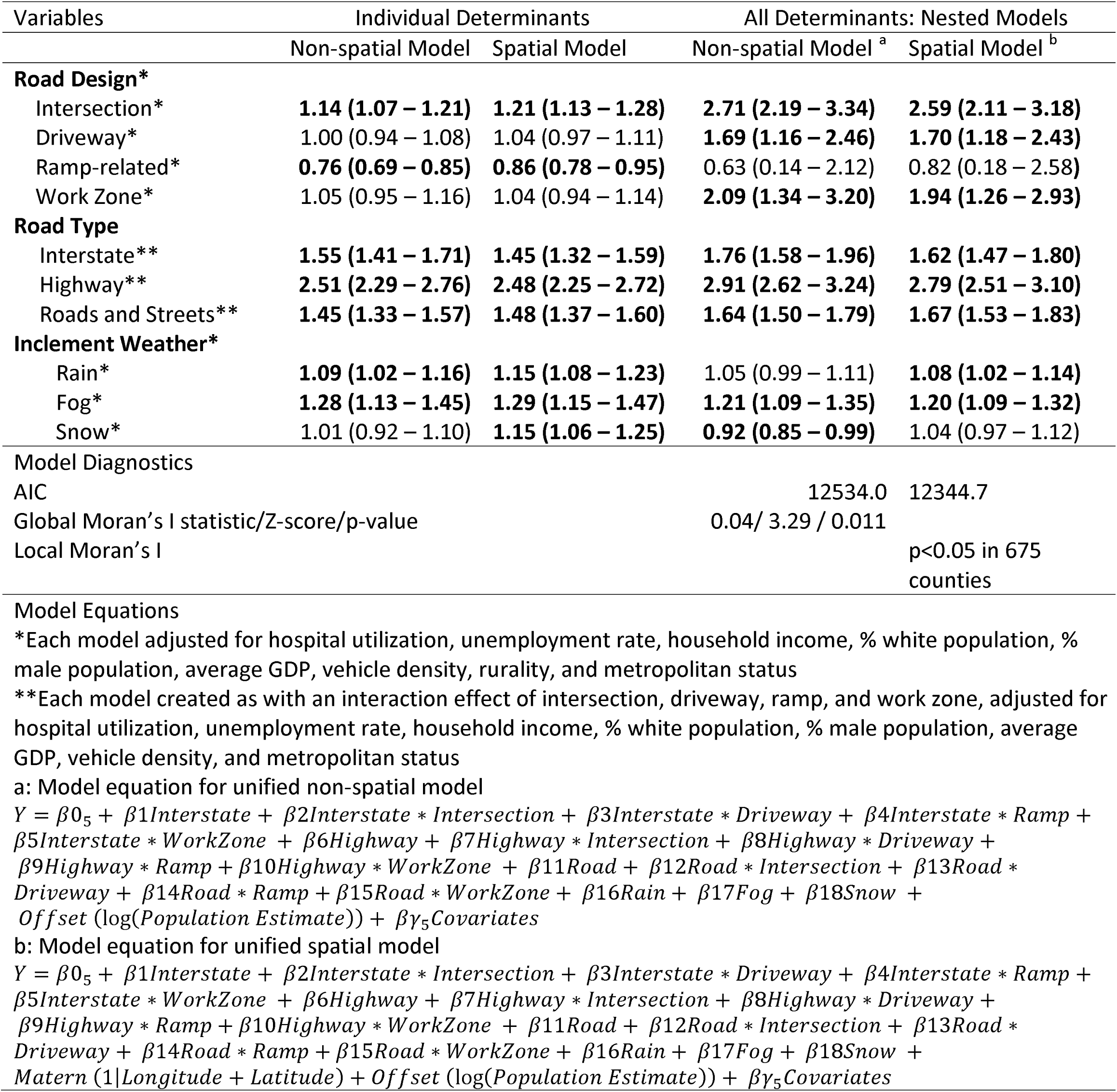
Negative binomial regression models predicting rush hour-related fatal road accidents occurring at road environmental and county-level characteristics.

In this study, the all-determinant spatial nested model performed better than the individual and non-spatial all-determinant models. The AIC of the all-determinant spatial model was lower than each of the individual determinant models (result not shown) and lower than the non-spatial all-determinant model (result not shown). Additionally, the significant Global Moran’s I of the residuals of the model suggested the presence of spatial autocorrelation (p=0.011) and further strengthened the need for a spatial model. Contrary to the earlier results, the spatial model showed that ramps were not protective against fatal crash injury, and the snow was not associated with increased fatal crash injury. Intersection (RR: 2.59; 95% CI: 2.11-3.18), driveway (RR: 1.70; 95% CI: 1.18-2.43), work zone (RR: 1.94; 95% CI: 1.26-2.93), interstate (RR: 1.62; 95% CI: 1.47-1.80), highway (RR: 2.79; 95% CI: 2.51-3.10), roads and streets (RR: 1.67; 95% CI: 1.53-1.83), rain (RR: 1.08; 95% CI: 1.02-1.14), and fog (RR: 1.20; 95% CI: 1.09-1.32) were associated with increased rate of fatal crash injury during the rush hour period.

### 4.3 Spatial Distribution

The crude fatal counts and small area estimates from the all-determinant spatial model for all the U.S. counties were displayed using choropleths maps (Figure 2). Twenty-two counties, located in California, Nevada, Arizona, Texas, Florida, Michigan, Illinois, and New York, had elevated crude and predicted rush hour-related fatal counts (>50 fatalities) (Figure 2B).

**Figure 2:**
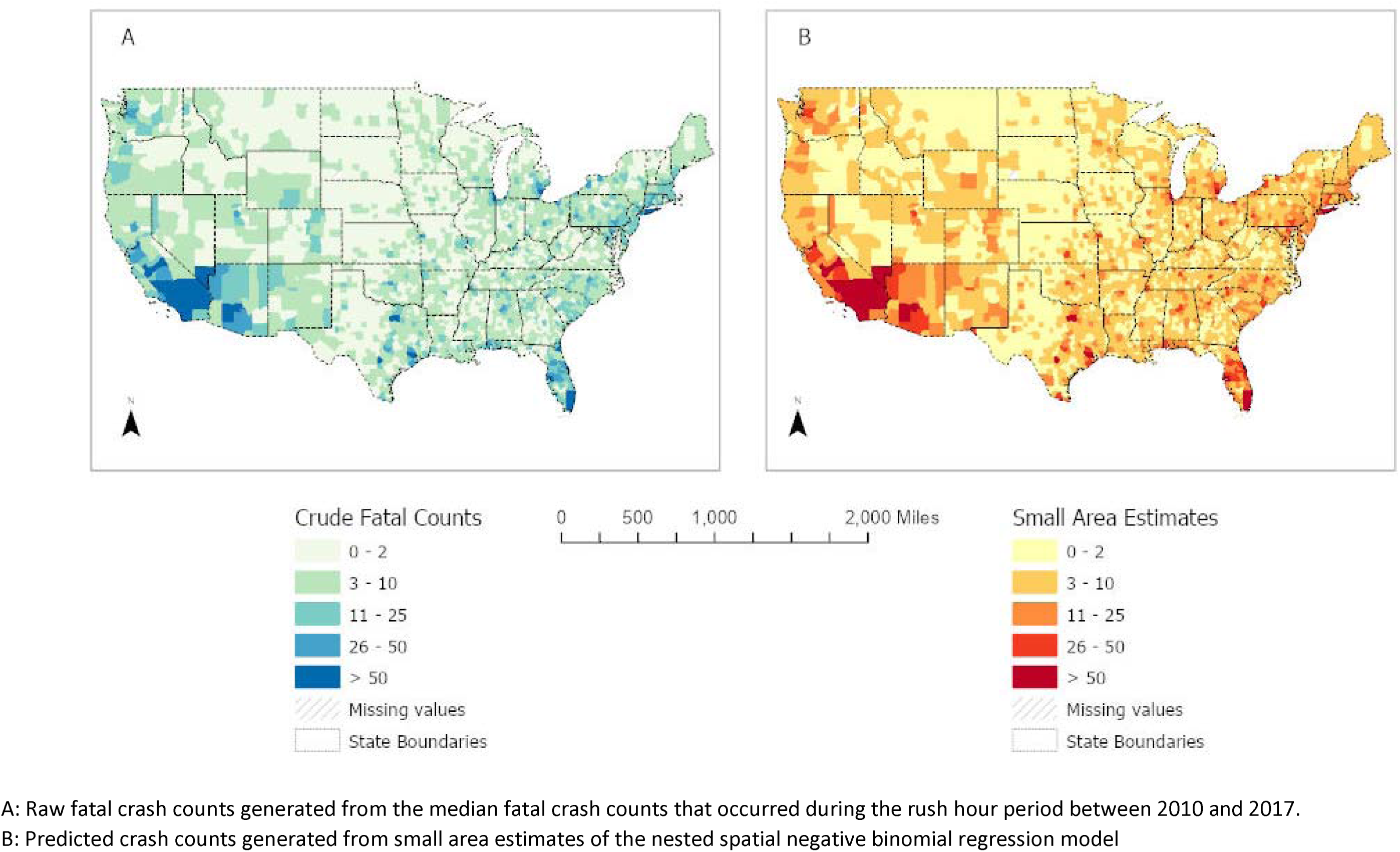
Raw (A) and Predicted (B) Median Rush-Hour Fatality Crash Counts per County: 2010 – 2017

We generated the crude and adjusted fatality rates using the average county population as the denominator (Figure 3). A total of 64 counties located in 22 states had crude fatality rates above 50 deaths per 100,000 population during the rush hour period (Figure 3A). However, after adjusting for county characteristics and the environmental determinants, only three counties, located in Kansas and Wyoming, had fatality rates in excess of 50 deaths per 100,000 population during the rush hour period (Figure 3B).

**Figure 3:**
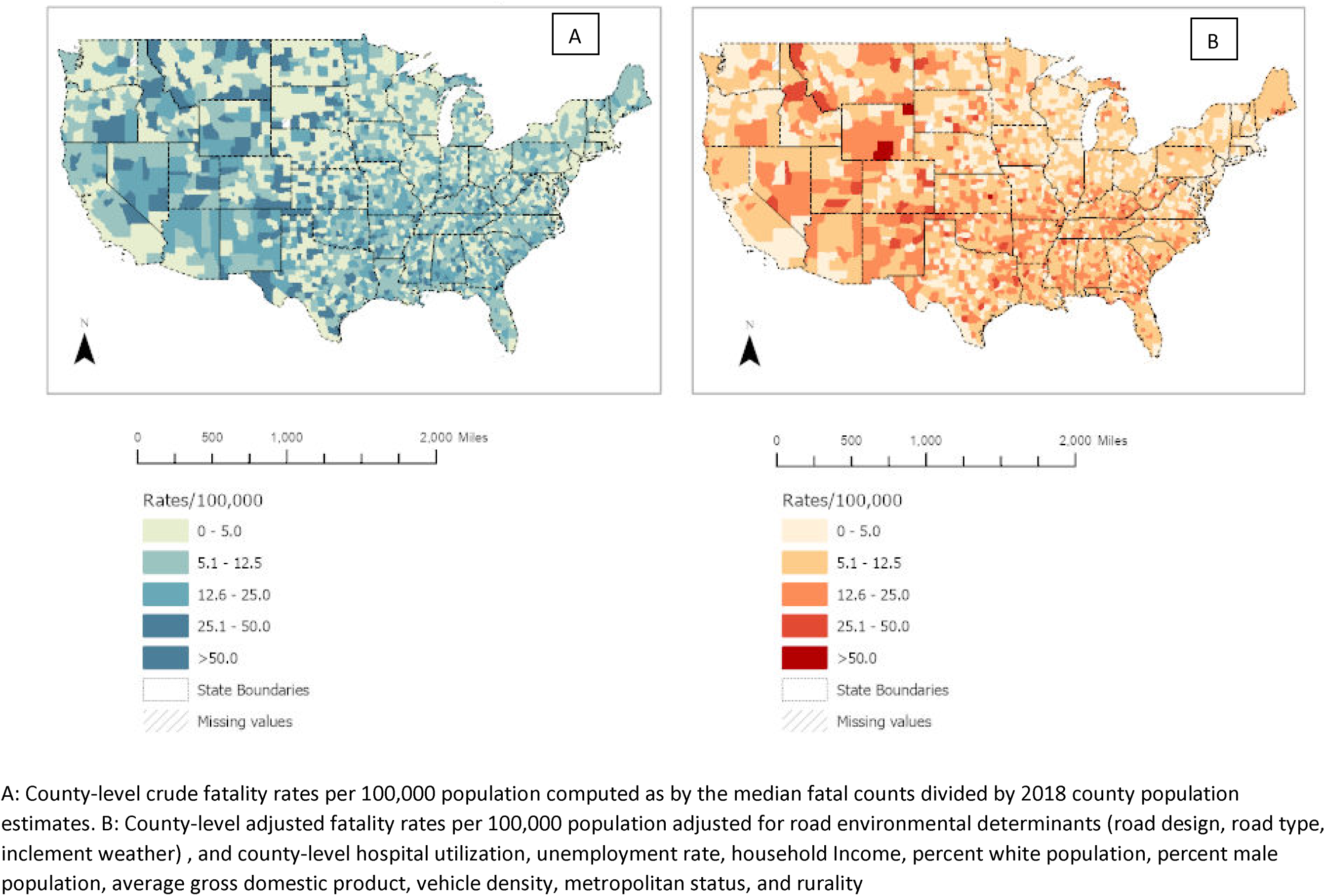
Crude (A) and Adjusted (B) Fatality Rate of Rush Hour related Fatal Crash Injury per County: 2010 – 2017

A cluster and outlier analysis showed that "high-high" clusters of rush hour-related fatal events in counties located in Idaho, Montana, Nevada, California, Wyoming, Utah, and across a few states in the Southeast (Figure 4A). Similarly, a hotspot analysis identified several counties in California, Nevada, Idaho, Montana, North Dakota, South Dakota, Wyoming, New Mexico, Colorado, and states in the Southeast as significant hotspots for rush hour-related fatal crash events (Figure 4B).

**Figure 4:**
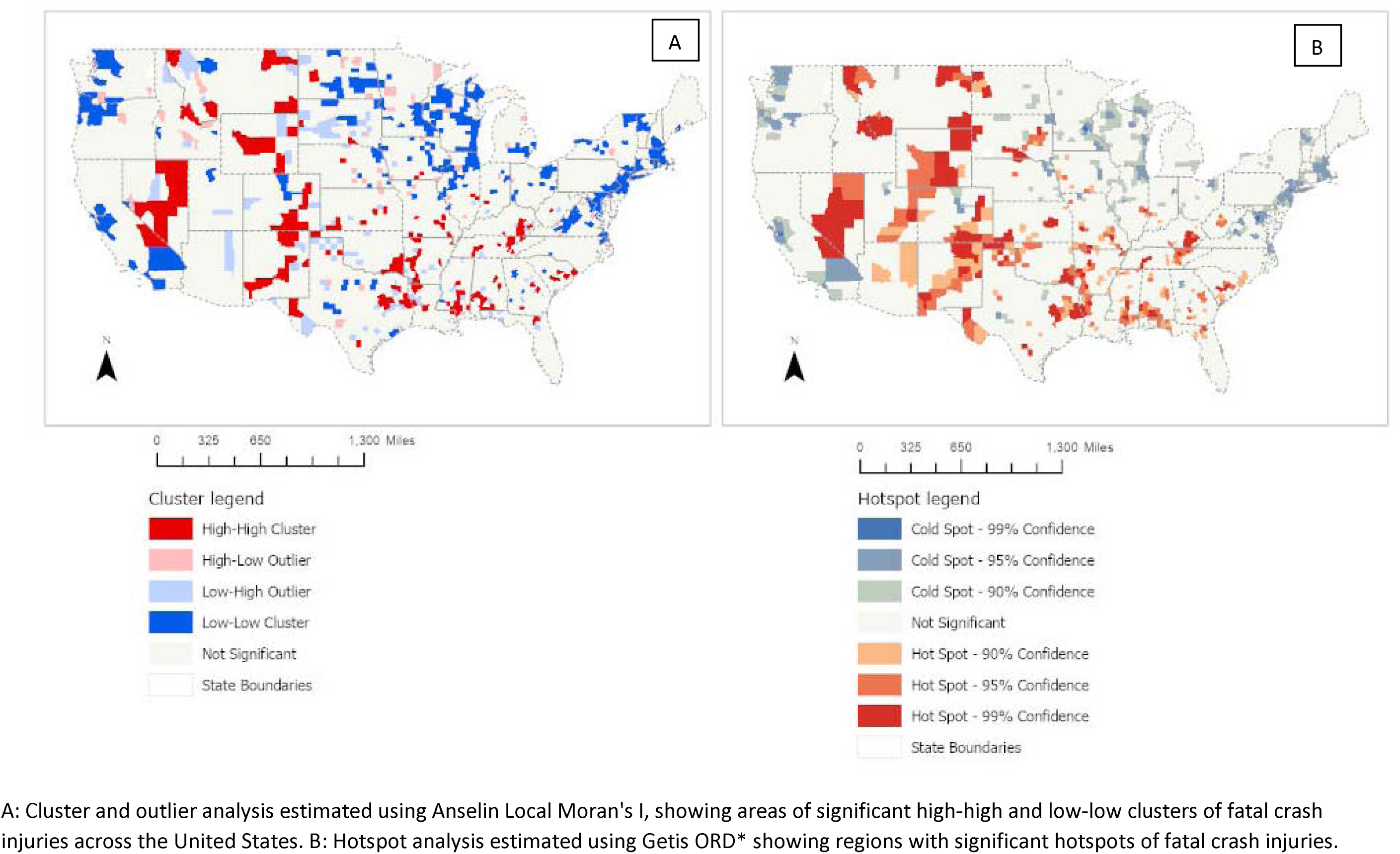
Cluster and Outlier analysis (A) and Hotspot Analysis (B) of Rush Hour-Related Fatal Crash Injuries per County: 2010 – 2017

## 5. Discussion

In this study, the prevalence of rush hour-related fatal crashes was 7.3 per 100,000 population. Case-specific fatality rates from interstates, highways, roads, streets, intersections, rain, fog, and snow were higher than the median fatality rates. Also, the median crash fatality rates were significantly higher in rural counties as compared to urban counties. During the rush hour period, fatal crash injury rates were significantly elevated on interstates, highways, roads and streets, intersections, driveways, and work zones. Further, rain and fog were significantly associated with fatal crash rates during the rush hour period. Rush hour-related fatal crash injuries disproportionately affected counties located in Idaho, Montana, Nevada, California, Wyoming, Utah, New Mexico, Texas, Colorado, Arkansas, Kentucky, Tennessee, and Alabama.

For over three decades, nationally representative crash reports have consistently reported elevated fatal crash counts in rural communities compared to urban communities (Insurance Institute for Highway Safety, 2019; National Center for Statistics and Analysis, 2017b; TRIP, 2020). It was not until 2016 that a reversal of trend showed an increasing crash count in urban communities with a subtle decline in rural communities (Insurance Institute for Highway Safety, 2019; National Center for Statistics and Analysis, 2017b). Earlier studies have attributed elevated rural fatal crash injuries to speeding (Insurance Institute for Highway Safety, 2019) and poor road conditions (Congressional Research Service, 2018; TRIP, 2020). Additionally, we report that some rural-urban socioeconomic differences exhibit significant associations with fatal crash injuries. In rural counties, increased White population proportion, vehicle density, median household income, and decreased male proportion are associated with reduced fatal crash injury during the rush hour period. Conversely, in urban counties, increased hospital utilization, unemployment rate, the proportion of Whites and males, county GDP, and decreased median household income were associated with increased fatal crash injury rates. These non-causal observatory findings identify how the social determinants of health differentially influence the fatal crash injury patterns in rural and urban environments (Healthy People, 2020).

In decreasing order of prevalence rates, road type-specific crash fatality rates were highest on the highways, followed by road and streets, and on interstates during the rush-hour period. The fatality rates pattern follows a similar pattern with the rates higher on highways, followed by roads and streets and interstates. The contrast in the prevalence rates on highways and on the interstate may be a reflection of the rush hour period. During the rush-hour period, highway road users are more likely to be residents within the state going from their homes to their workplaces in the morning and vice-versa in the evening. Contrastingly, interstate road users may be traversing different counties and states, although some workers engage in long commutes to work (Di Milia, Rogers, & Åkerstedt, 2012). Further, the interstate accounts for less than 2% of the total road mileage on all U.S. roads, but about 24% of all travel occurs on the interstate (Federal Highway Administration, 2000a). Irrespective of the road type, travel duration, and mileage are associated with increased fatality rate (Rolison & Moutari, 2018).

We report an elevated rate of fatal crash injuries at intersections, driveways, and work zones during the rush hour period. With more than 50% of fatal and non-fatal crashes occurring at intersections (Federal Highway Administration, 2020b), it was expected that the intersection-specific fatality rate would be higher than driveway, ramp, and work zone-specific fatality rates. Speeding and driving inattention might be some of the reasons associated with increased fatal crash rate at intersections, driveways, and work zones. Liu et al. (2007) reported that rush-hour driving was associated with the speed at which drivers approach the intersections. NHTSA reported that misjudgment of another vehicle’s speed and inadequate surveillance was associated with four to six-fold increased odds of fatal crash injuries (National Highway Traffic Safety Administration, 2010). A recent meta-analysis reported that increasing work zone driving duration increases crash rates by approximately three folds, and for every kilometer increase in the length of the work zone region, the crash rate increases by two folds (Athanasios Theofilatos et al., 2017).

Earlier studies have reported that rain-related fatal crash injuries account for 8-10% of all fatal crash counts (Black, Villarini, & Mote, 2017; S. Saha, Schramm, Nolan, & Hess, 2016). In addition, we report elevated rates of fatal crash injuries from rain, with the rates significantly higher in rural counties during the rush hour period. However, fog-specific fatal crash rates were higher than rain and snow-specific fatal crash rates during the rush hour. Additionally, the rate of fog-related fatal crashes was higher than the rate associated with rain during the rush hour period. The increased fatality rate from these adverse weather events may be associated with decreased visibility (El-Basyouny et al., 2014; Li et al., 2019). Earlier studies have reported reduced speeding when driving in the rain, snow, and fog (Federal Highway Administration, 2020a; Y. N. Miller, Hilpert, Klein, Tyler, & Brooks, 2007; Wu et al., 2018).

Despite the increased rate of fatal events at different road types and road designs, the rates are disproportionately distributed across the U.S. This study and earlier studies (Byrne et al., 2019; National Center for Statistics and Analysis, 2019a; National Highway Traffic Safety Administration, 2016b) have reported worse crash rates and increased rate of fatal crash injuries in rural counties as compared to urban counties. However, identifying counties in need of focused crash interventions, especially during the rush hour, may hold the solution to achieving zero fatality rates. We demonstrated that counties located in states identified in this study serve as clusters and hotspots for fatal crash events during the rush hour after adjusting for environmental and county characteristics. Earlier studies have reported increased crash fatality rates in similar states (Ecola et al., 2018; National Highway Traffic Safety Administration, 2018).

Prioritizing intervention by states is not a novel approach to reducing fatal crash injury. There have been reports urging state-specific interventions towards specific risk factors associated with fatal crash events (Ecola et al., 2015; Ecola et al., 2018). Since each state within the U.S. is responsible for enacting policies and implementing crash prevention programs (Ecola et al., 2015), the need for tools that will guide decisions and policymakers on prioritization is needed. The Motor Vehicle Prioritizing Interventions and Cost Calculator for States (Center for Disease Control and Prevention, 2020b) represent one of those decision tools, which focuses primarily on risky driving behavioral intervention. This study demonstrates the need for a complementary tool that will help each state improve the road environmental network.

This study compared estimates from the nested spatial negative binomial model and the non-spatial model. The spatial model performed better, evidenced by the model diagnostic information. Further, this study highlights the benefits of perfunctorily assessing spatial autocorrelation as the use of spatial estimators influences the results of the study. An argument may be made on how much improvement the spatial model provides. The small value of the global Moran’s I may be a reflection of long-range dependencies and the decay with increasing distance (Epperson, 2005). We demonstrate that the spatial model improved the model and produced marginally better estimates with narrower confidence intervals. For example, the nested spatial model showed that there was no significant relationship between snow and fatal crash injury while the non-spatial model would have been falsely interpreted as snow being associated with reduced odds of fatal crash injury during the rush hour period.

Predicting crash injury rate ratios from environmental characteristics requires establishing a hierarchical modeling approach (Alarifi, Abdel-Aty, & Lee, 2018; D. Saha, Alluri, Gan, & Wu, 2018) or nested (Abdel-Aty & Abdelwahab, 2004; Patil, Geedipally, & Lord, 2012). For this study, a nested model was intuitive as road designs are contained within each road type. Earlier studies have adopted other spatial modeling methodologies such as geographically weighted Poisson regression models (Bao, Liu, & Ukkusuri, 2019; Goldstick, Carter, Almani, Brines, & Shope, 2019; Hezaveh, Arvin, & Cherry, 2019), ordered probit model (Castro, Paleti, & Bhat, 2013), spatial autoregressive model (Dezman et al., 2016), multiple additive Poisson regression models (Ding, Chen, & Jiao, 2018), and multivariate Poisson lognormal spatial model (Jonathan et al., 2016). These models were influenced mainly by how crash injury was defined and the choice of predictor variables. This study, which used a nested model, adds to the crash injury literature an additional prediction model. We demonstrate its parsimonious use in assessing environmental characteristics, using the lens of social determinants of health.

This study has its limitations. Because of its ecological nature, causal inferences cannot be established. FARS dataset relies on crash reporting across all states. Therefore, data entry and processing errors cannot be eliminated. The rush-hour period varies widely across states and counties. Therefore, our definition of the rush-hour period may overestimate the rush-hour period in some counties and underestimate others. Due to the non-static traffic pattern across counties, there is a possibility of misclassification bias. However, such misclassification is likely to be non-differential. Despite these limitations, this study is one of the few studies that identify regions requiring crash injury-related interventions. The national, rural, and urban estimates of the average median fatal crash injury rates fill the gap in the crash injury prevention literature; there is no recent study that quantified rush hour-specific crash prevalence and fatality risks. Additionally, this study provides information that can inform policy and resource allocation in the presence of other competing public health issues.

## 6. Conclusion

As the U.S. journeys toward achieving a zero-fatal crash injury rate, identifying the built and natural environmental elements associated with fatal crash injuries can inform policy and practice. Also, understanding the rural and urban differences in fatal crash injury patterns during the rush hour period and counties and clusters where significant injuries occur will help identify areas needing focused intervention. This study identifies road environmental characteristics associated with fatal crash injury during the rush hour period and demonstrates, through spatial modeling tools, areas that may need focused intervention. While this study provides information on the rush hour-related crashed, an understudied domain in crash injury prevention, it provides a useful tool that can guide policy, public health practice, and resource allocation at the national, state, and county levels.

## Data Availability

Data is publicly available and freely downloadable at the National Highway Traffic Safety Administration website

https://www.nhtsa.gov/file-downloads?p=nhtsa/downloads/FARS/

